# In vitro splice-switching oligonucleotide rescues aberrant *GFM2* pseudoexon inclusion and restores mitochondrial activity

**DOI:** 10.64898/2026.05.28.26354078

**Authors:** Shira Gross, Rivka Birnbaum, Nava Shaul Lotan, Hagar Mor-Shaked, Joshua Manor, Avraham Shaag, Chaggai Rosenbluh, Adi Levy-Mamo, Shira Yanovsky-Dagan, Ann Saada, Tamar Harel

## Abstract

**Background:** Biallelic variants in *GFM2,* encoding mitochondrial elongation factor G2 (mtEFG2), a GTPase involved in the termination stage of mitochondrial translation, cause autosomal recessive combined oxidative phosphorylation deficiency. Noncoding structural variants may be missed by exome sequencing but can disrupt splicing and provide opportunities for variant-specific therapeutic rescue. We investigated the molecular mechanism underlying suspected Leigh syndrome in an infant with mitochondrial disease and evaluated whether splice-switching oligonucleotide (SSO) treatment could correct the pathogenic splicing defect.

**Methods:** The proband underwent exome sequencing followed by short-read and long-read whole genome sequencing. RNA sequencing, reverse-transcription PCR, quantitative PCR, and cycloheximide treatment were used to characterize the effect of the identified intronic duplication on *GFM2* splicing and transcript stability. Patient-derived fibroblasts were treated with SSOs targeting the aberrant splice junction. Rescue was assessed by RNA studies, western blotting, and spectrophotometric measurement of cytochrome c oxidase (COX).

**Results:** Whole genome sequencing identified a paternally-inherited *GFM2* missense variant, NM_032380.5:c.2195C>T p.(Pro732Leu), in trans to a maternally-inherited 221-nucleotide intronic duplication, NM_032380.5:c.2029-741_2029-521dup. RNA studies revealed a 87-nucleotide pseudoexon, generated by activation of a cryptic acceptor splice site within the duplicated sequence. The resulting transcript harbored a premature termination codon (PTC) and underwent nonsense-mediated decay, as confirmed by cycloheximide rescue. Together with reduced mtEFG2 protein levels on western blot, the findings supported a loss-of-function mechanism. Enzymatic analysis of affected fibroblasts showed reduced activity of the mtDNA-dependent complex IV subunit COX, with preservation of the nuclear-encoded complex II enzyme succinate dehydrogenase and the control enzyme citrate synthase, consistent with impaired mitochondrial translation. A SSO targeting the aberrant intron-pseudoexon junction nearly abolished pseudoexon inclusion, restored correctly spliced *GFM2* transcript from the duplication-containing allele, increased mtEFG2 protein levels, and significantly improved COX activity.

**Conclusions:** This study identifies a pathogenic intronic *GFM2* duplication that causes mitochondrial disease through pseudoexon activation and nonsense-mediated decay. The findings demonstrate the value of integrated genome and transcriptome analysis for exome-negative mitochondrial disease and provide in-vitro proof of concept that SSOs can restore transcript processing, protein expression, and mitochondrial respiratory-chain function in patient-derived cells.

## BACKGROUND

Mitochondrial diseases are a diverse group of inherited disorders that arise from disruptions in the process of oxidative phosphorylation (OXPHOS), which is the principal pathway for adenosine triphosphate (ATP) synthesis (1–3). Impairment of this pathway can cause energy shortages, particularly in high-energy demand tissues, including the brain, peripheral nerves, eye, heart, and skeletal muscle. Accordingly, affected individuals may present with developmental delay, seizures or encephalopathy, hypotonia or myopathy, cardiomyopathy, ophthalmologic disease, or multisystem involvement. Since approximately 340 nuclear and mitochondrial genes contribute to mitochondrial function, these disorders show marked clinical and molecular variability (4–6).

The OXPHOS system consists of five major multi-subunit complexes: complexes I to IV, which form the electron transport chain (ETC), and complex V (ATP synthase). Together, these complexes generate the vast majority of cellular energy in the form of ATP. The OXPHOS system comprises over 90 subunits of which only thirteen are encoded by mitochondrial DNA (mtDNA). With the exception of complex II, which is entirely nuclear-encoded, OXPHOS complexes contain both nuclear- and mitochondrial DNA (mtDNA)-encoded subunits, reflecting bi-genomic regulation. Additional nuclear genes encode proteins involved in mtDNA maintenance, as well as mitochondrial transcription, translation, and other essential mitochondrial functions. Thus, pathogenic variants affecting either nuclear or mitochondrial genes can impair ATP production and cause mitochondrial disease (7, 8).

Combined oxidative phosphorylation deficiencies (COXPD) are most often caused by pathogenic variants in mtDNA-encoded mitochondrial rRNAs and tRNAs or by nuclear gene defects affecting mitochondrial protein synthesis, mtDNA maintenance, or assembly of the respiratory chain complexes. Leigh syndrome is one of the most severe pediatric presentations of mitochondrial energy failure (9, 10). It typically presents in infancy or early childhood and is characterized by neurodevelopmental impairment, psychomotor regression, feeding difficulties, dystonia, seizures, respiratory compromise, and, in some individuals, cardiomyopathy or renal involvement. Brain magnetic resonance imaging (MRI) most often demonstrates abnormalities of the basal ganglia and brainstem. Clinically, Leigh syndrome manifests with developmental delay, psychomotor regression, feeding difficulties, dystonia, seizures, respiratory problems, and sometimes involvement of other organs such as cardiomyopathy or renal failure. Brain MRI classically demonstrates symmetric lesions of the basal ganglia and/or brainstem (11). The genetic heterogeneity of Leigh syndrome and related COXPDs makes a precise molecular diagnosis important for prognosis, reproductive counseling, and emerging mechanism-based therapeutic strategies (12).

Pathogenic variants in genes encoding components of the mitochondrial translation machinery, including *GFM1* and *GFM2* cause specific forms of COXPD (13–17). *GFM2*, located on chromosome 5q13.3, encodes mitochondrial elongation factor G2 (mtEFG2) which functions during the ribosome-recycling stage that follows mitochondrial translation termination (13). In contrast to the bacterial EF-G, which handles multiple functions, human mitochondria use two specialized homologs. mtEFG1, encoded by *GFM1*, mediates mitoribosomal translocation during elongation, whereas mtEFG2 is recruited after polypeptide release and acts with the mitochondrial recycling factor mtRRF to promote dissociation of the mitoribosome into subunits (18). The recycling process is tightly regulated and is required for mitochondrial translation and efficient oxidative ATP production (19, 20). Defects in any part of this pathway, including *GFM2* variants, lead to reduced synthesis of mtDNA-encoded OXPHOX subunits and thereby impair mitochondrial respiration, causing severe mitochondrial diseases with early-onset neurologic symptoms (13).

Splice-switching oligonucleotides (SSOs) are short synthetic antisense molecules that bind specific sequences in pre-mRNA and alter splice-site recognition without necessarily inducing RNA degradation. By blocking cryptic splice sites, splice enhancers, or other cis-regulatory elements, SSOs can restore normal splicing, promote exon skipping, or modulate exon inclusion. This sequence-specific mechanism has particular relevance for rare genetic disorders caused by pseudoexon activation or other variant-specific splicing defects (21, 22).

Here, we present an infant with suspected Leigh syndrome caused by compound heterozygous *GFM2* variants detected by whole genome sequencing (WGS): a missense variant in *trans* to an intronic duplication that activates pseudoexon inclusion. Using DNA, RNA, protein, and biochemical studies, we establish the pathogenic mechanism and show that an SSO designed to block the aberrant splice junction rescues normal *GFM2* splicing, increases mtEFG2 protein, and improves mitochondrial respiratory-chain activity in affected fibroblasts.

## METHODS

### Exome sequencing

Following informed consent, genomic DNA was extracted from peripheral blood. Exome enrichment was performed using the IDT xGen Exome Research Panel v2.0 (Integrated DNA Technologies, IDT), followed by 100-base pair (bp) paired-end sequencing on an Illumina NovaSeq 6000 platform (Illumina, San Diego, California, USA).

FASTQ filess were uploaded to the Geneyx Analysis platform (23). Alignment and variant calling for single nucleotide variations (SNVs) and copy number variants (CNVs) were performed using Illumina DRAGEN Bio-IT. VCF files were annotated with the Geneyx Analysis annotation engine, and reviewed for analysis, filtering and interpretation. The human genome assembly GRCh38 was used as reference. Variants were excluded if they were off-target (intronic variants >48 bp from a splice junction), synonymous unless located within 54 bp of a splice site, or present at minor allele frequency (MAF) >0.01 in gnomAD or our internal database.

### Short-read WGS

Genomic DNA was extracted from peripheral blood and next-generation sequencing libraries were prepared using the Illumina PCR-free TruSeq DNA Library Prep Kit. Sequences were generated on an Illumina NovaSeq 6000 sequencing platform as 150-bp paired-end reads, to a final depth of 73X. FASTQ files were uploaded to the Geneyx Analysis platform (23). Alignment and variant calling for SNVs, CNVs, structural variants, and tandem repeats were performed using Illumina DRAGEN Bio-IT. VCF files were annotated on the Geneyx Analysis annotation engine, and presented for analysis, filtering and interpretation, using GRCh37/hg19 as the reference genome. Variants were manually lifted over to GRCh38/hg38.

### Long-read WGS

Genomic DNA quality was assessed using a Nanodrop and the Qubit dsDNA HS Assay Kit (Thermo Scientific Fluoroskan). DNA was mechanically fragmented with FastPrep-96 and normalized to 1–2 ng in 50 µL. Libraries were prepared using the SQK-LSK-114 kit (Oxford Nanopore Technologies, ONT) and the NEBNext® Companion Module for ONT (E7180S), loaded onto R10.4 flow cells, and sequenced on a PromethION48 for 72h with high-accuracy base calling and 5mC methylation detection (ONT, FLO-PRO002). Base calling was performed with Guppy, reads were aligned with minimap2, and SNVs and structural variants (SVs) were called with Clair3 (SNVs) and Sniffles2 (SVs). VCF files were annotated with the Geneyx Analysis platform using GRCh38 as the reference genome.

### RNA sequencing

Total RNA was extracted from peripheral blood using the QIAamp RNA Blood Mini Kit (Qiagen, Hilden, Germany), and poly(A)-selected sequencing libraries were prepared with the KAPA RNA HyperPrep kit (Roche, Basel, Switzerland), according to the manufacturers’ protocols. cDNA libraries were sequenced on an Illumina NovaSeq6000 sequencing platform with 161-bp paired-end reads. RNA-seq reads were aligned to the hg19 reference genome with STAR 2.7.9 using an index optimized for 161-bp read length and the two-pass Basic mode.

### cDNA analysis

Total RNA was extracted from fibroblasts of affected and control individuals, using TRIzol reagent. First-strand cDNA was synthesized from 1 μg total RNA with the qScript cDNA Synthesis Kit (Quantabio), which combines oligo(dT) and random primers. Regions spanning the relevant exons were amplified using PCRBIO HS Taq Mix Red (PCR biosystems, London, UK); primers are provided in Table S1. The resultant fragments were separated by 1.5% (w/v) agarose gel electrophoresis and analyzed by Sanger sequencing.

### Breakpoint junction analysis

The duplication breakpoint was confirmed at the DNA level, using primers F_GFM2_int19 and R_GFM2_int19; and at the RNA level, using primers F_GFM2_Jx18-19 and R_GFM2_Ex20 (Table S1, Fig. S1). PCR products were separated by 1.5% agarose gel electrophoresis and analyzed by Sanger sequencing.

### Quantitative PCR (qPCR)

qPCR was performed to quantify transcript levels using gene-specific primers and SYBR dye on a real-time PCR system. Reactions were performed on triplicate experiments, each with technical triplicates, and relative expression was calculated using the comparative Ct (ΔΔCt) method after normalization to a housekeeping gene. Primers were designed to amplify total *GFM2* transcript (F_GFM2_Ex16 and R_GFM2_Ex17) or the pseudoexon-containing transcript (F_GFM2_Jx19-PE and R_GFM2_Ex21). Primer design and sequences are provided in Table S1 and Fig. S1.

### Tissue culture

Primary fibroblasts derived from a skin biopsy of the proband were maintained in Dulbecco’s Modified Eagle’s Medium (DMEM) supplemented with 15% fetal calf serum, 1% L-glutamine, and 1% penicillin-streptomycin (Biological Industries Beit Haemek, Israel). Primary fibroblasts were immortalized with a lentivirus expressing human telomerase reverse transcriptase (hTERT) that was generated by transfecting HEK293T cells (ATCC) with the plasmids pLV-UBC-hTERT (Addgene no. 85140), psPax2 (Addgene no. 12260), and pMD2.G (Addgene no. 12259). Cells were selected with blasticidin (10 μg/mL; Bio Prep). hTERT-immortalized fibroblasts were treated with 100µg/ml Cycloheximide (CHX) for 24 hours, followed by RNA extraction from treated and untreated cells.

### Western blot

Cells were lysed in RIPA buffer (R22041DV) supplemented with protease inhibitors, and protein concentrations were determined by Bradford assay. Equal amounts of total protein were separated by SDS–PAGE and transferred to PVDF membranes. Membranes were blocked with 5% skim milk and incubated with primary antibodies against total GFM2 (anti-GFM2, 1:1000) (Proteintech, 16941-1-AP), followed by incubation with appropriate HRP-conjugated secondary antibody. Bands were detected by chemiluminescence, quantified using ImageJ software, and normalized to a loading control.

### Enzymatic activity measurements

Enzymatic activities of cytochrome c oxidase (COX), succinate dehydrogenase (SDH) and citrate synthase (CS), were measured in cell sonicated homogenates using spectrophotometric methods as previously described.(24) COX activity was determined by monitoring the oxidation of reduced cytochrome c at 550 nm. CS activity was assessed in the presence of acetyl-CoA and oxaloacetate by quantifying the coenzyme A (CoA-SH) released during the reaction, which forms a colored complex with 5,5′-dithiobis-(2-nitrobenzoic acid) (DTNB) at 412 nm. SDH activity was measured by monitoring the decrease in 2,6 dichoroindophenol (DCIP) absorbance at 600 nm in presence of succinate and phenazine methosulfate (PMS). Experiments were done on three independent samples, each with technical triplicates. Measurements were performed on a Uvikon double beam spectrophotometer (Kontron Instruments Ltd, USA). Statistical analyses were performed using GraphPadPrism v10 (GraphPad Software, San Diego, CA, USA). Group comparisons were conducted by analysis of variance (ANOVA).

### Splice-switching oligonucleotides

Two SSOs (Table S2) were designed to target the 5’ and 3’ exon-intron junctions based on Garanto and Collin (2018) (25), considering length, GC content, melting temperature, secondary structure and free energy of the SSO, secondary structure of the target region, binding energy, and sequence uniqueness. SSOs were ordered from IDT with 2’-O-methyl (2’OMe) modifications and a phosphorothioate backbone. A sequence-scrambled SSO of equivalent length and modification was used as a negative control (Table S2). Affected and control fibroblasts were seeded in 6 well plates on day 0 and transfected on day 1 using 7.5 μl RNAiMAX reagent (Invitrogen) and either 30 or 150 pM SSO in 500 µL Opti-MEM, followed by addition of 2 mL medium. RNA and protein were extracted 48 hours after transfection and pellets for functional assays were harvested 72 hours after transfection.

## RESULTS

### Clinical Report

The proband (Fig. 1A, Individual II-2) was a young boy who presented with seizures. He was the son of a consanguineous couple with one older healthy sister. Proband was born at term with a birthweight appropriate for gestational age, following a pregnancy notable for reduced fetal movements. Early concerns included poor eye contact, feeding difficulties, irritability, failure to thrive and global developmental delay, with no developmental milestones achieved either at presentation or upon reassessment. Additional clinical features included hypospadias and a history of surgical correction for undescended testis and inguinal hernia.

**Figure 1.**
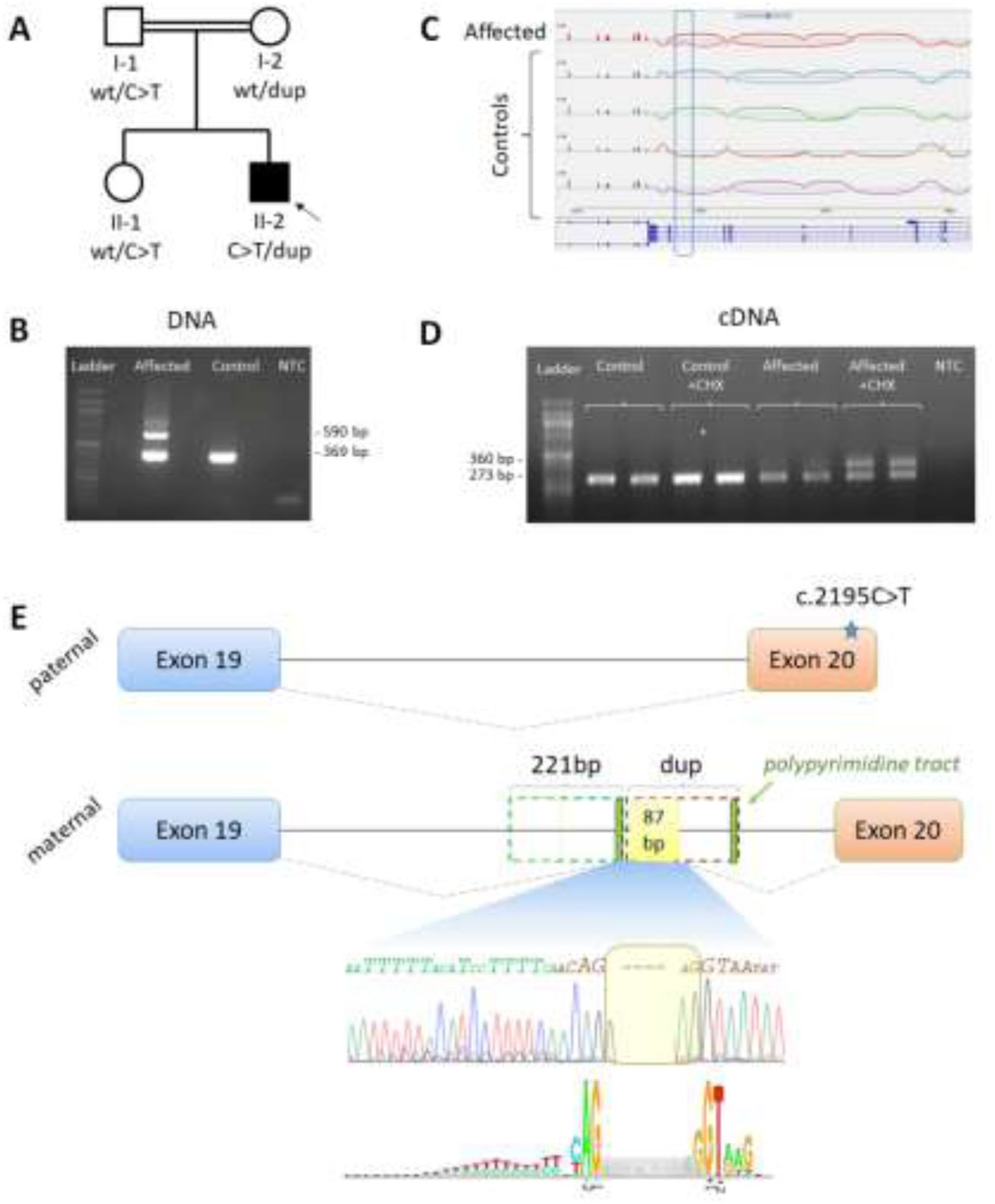
Genomic and transcript-level characterization of a *GFM2* intronic duplication causing pseudoexon inclusion. (A) Pedigree demonstrating familial segregation of the variants in the proband and relatives. (B) Agarose gel electrophoresis of PCR products amplified from genomic DNA of affected and control individuals. (C) Sashimi plot of blood RNA sequencing from the affected and controls, showing additional exon inclusion in the affected individual’s *GFM2* transcript. (D) PCR amplification of cDNA from affected and control fibroblasts, with and without cycloheximide (CHX) treatment; each sample was analyzed in duplicate. (E) Schematic representation of *GFM2* showing the paternal allele with the missense variant in exon 20 and a detailed view of the maternal allele containing the intronic duplication responsible for pseudoexon formation. The expanded sequence view highlights the novel splice junction generated by the duplication and its similarity to canonical splice-site motifs. The bold nucleotides indicate those resembling the consensus motif.

On examination, he displayed mildly dysmorphic facial features, a myopathic appearance, persistent lack of eye contact, axial hypotonia, and increased peripheral tone during crying. Laboratory analysis revealed elevated blood lactate (4.1 mmol/L) and CSF lactate (3.6 mmol/L), both exceeding the normal range (<2.1 mmol/L). Brain MRI demonstrated bilateral high signal intensity in the putamen with age-appropriate myelination. Metabolic work up, including plasma amino acids, urine organic acids, and biotinidase activity, was nondiagnostic.

Leigh Syndrome was suspected on the basis of the clinical, biochemical, and neuroimaging findings. Exome sequencing was unrevealing, prompting subsequent WGS for a more comprehensive analysis.

### WGS identifies an intronic duplication in *trans* to a missense variant in *GFM2*

WGS identified two variants in *GFM2*: a missense variant in exon 20 (chr5:74722395 G>A [hg38]; NM_032380.5:c.2195C>T p.(Pro732Leu)) inherited from the father and a 221-bp intronic duplication within intron 19 (chr5:74,723,083-74,723,303 [hg38]; NM_032380.5:c.2029-741_2029-521dup) inherited from the mother (Fig. 1A). PCR and Sanger sequencing confirmed the duplication (Fig. 1B) and demonstrated compound heterozygosity in the proband. The unaffected sister was heterozygous for the missense variant only (Fig. 1A). Long-read WGS confirmed the duplication breakpoint (Fig. S2, S3). Biallelic *GFM2* disruption was consistent with the proband’s presentation, prompting evaluation of the pathogenic potential of these variants. A review of the clinical features of reported individuals and the proband is provided as Table S3. The missense variant c.2195C>T was absent from gnomAD and had supportive *in silico* evidence for deleteriousness, including a CADD score of 29, GERP score of 5.96, AlphaMissense score of 0.779 and REVEL score of 0.91, meeting ACMG evidence codes PM2 and PP3_moderate (26).

### RNA studies identify pseudoexon inclusion

To assess the intronic duplication, wild-type and duplicated sequences were evaluated by splicing-prediction software (ASSP; http://wangcomputing.com/assp/), which predicted a high-scoring predicted constitutive acceptor splice site within the duplicated sequence (ASSP score: 8.778). RNA sequencing from blood (Fig. 1C) and reverse transcriptase PCR (RT-PCR) from fibroblast cDNA (Fig. 1D) both demonstrated an 87-nucleotide insertion in the *GFM2* transcript immediately downstream of the duplication junction, consistent with pseudoexon inclusion.

In fibroblasts, the insertion-containing band was markedly weaker than the wild-type band, suggesting degradation by nonsense-mediated decay (NMD). To investigate this, fibroblast cultures were treated with CHX. Following treatment, RT-PCR demonstrated a clear increase in the transcript containing the insertion (Fig. 1D), supporting that the aberrant transcript undergoes NMD. A modest increase in *GFM2* RNA was also observed in control cells after CHX treatment, consistent with the broader role of NMD in regulating physiological transcripts as well as mutant mRNAs (27).

Inspection of the duplicated DNA sequence in conjunction with the RNA data indicated that the duplication positioned a polypyrimidine tract directly upstream of a cryptic ’AG’ acceptor splice site, thereby activating pseudoexon inclusion (Fig. 1E).

### SSO prevents pseudoexon inclusion

Given that the intronic duplication activated a deleterious pseudoexon, we designed two SSOs to block the aberrant splicing event (Table S2). SSO1 targeted the cryptic splice junction created by the intronic duplication, whereas SSO2 targeted a predicted SC35/SRSF2 regulatory region near the 3’-end of the pseudoexon (Fig. 2A). The SC35/SRSF2-recognized exonic splicing enhancer motifs can promote spliceosome assembly and exon inclusion at nearby splice sites.^14^ Both SSOs were designed to act through steric blockade at the pre-mRNA level and carried 2′O-methyl (2′OMe) and phosphorothioate modifications to improve stability and activity (25).

**Figure 2.**
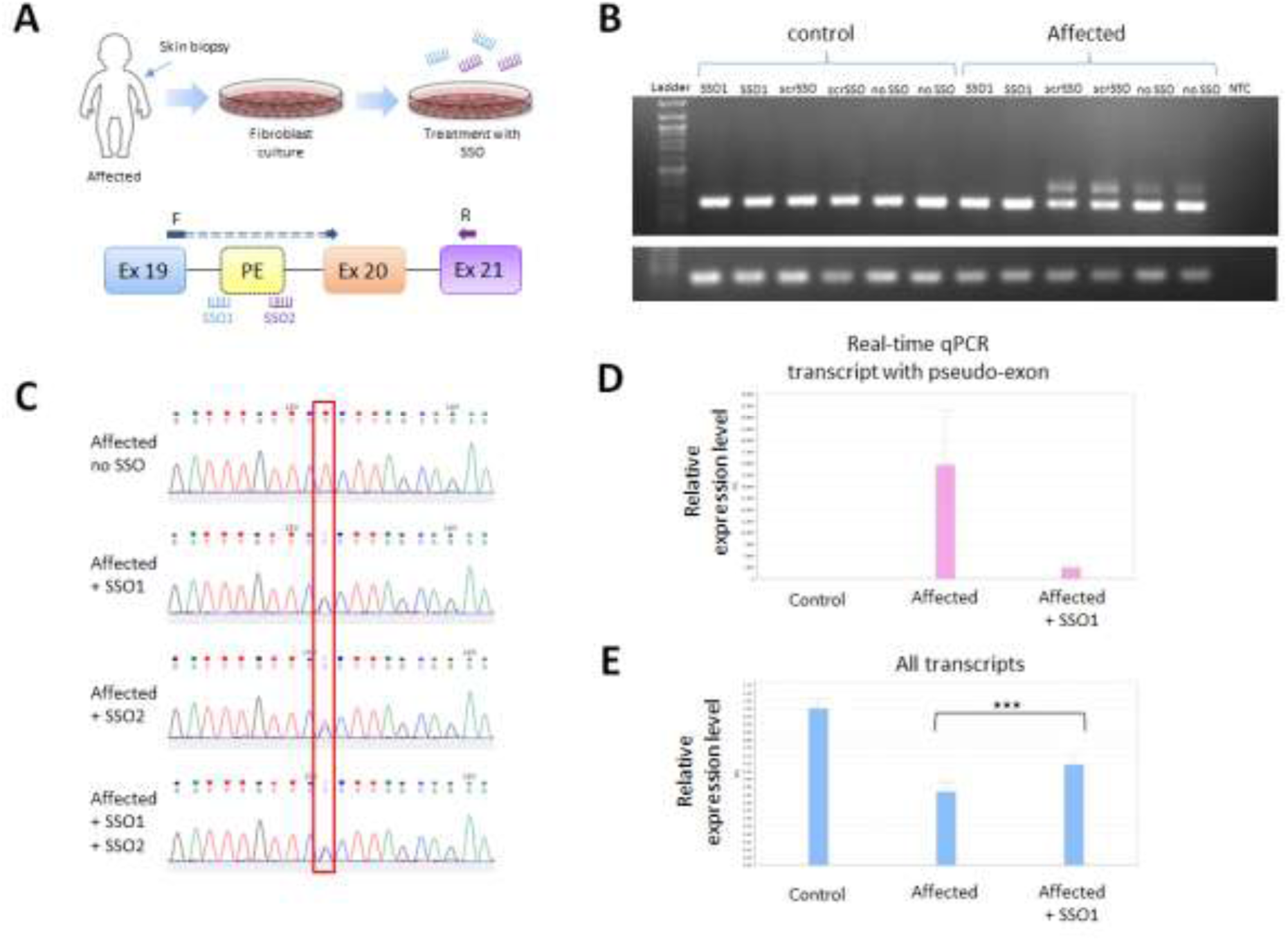
SSO-mediated correction of aberrant splicing and restoration of mitochondrial function in GFM2 deficient fibroblasts. (A) Schematic overview of fibroblast derivation from affected individual’s skin biopsy and subsequent SSO transfection. Schematic representation of the maternal *GFM2* transcript following splice-switching antisense oligonucleotides (SSO) treatment, with illustration of the primers that were used to specifically amplify the wild-type transcript. (B) PCR analysis of cDNA synthesized from SSO-transfected fibroblasts (150 pM) using primers amplifying both *GFM2* transcript isoforms. Lower panel shows *RPLPO* expression as a housekeeping control. (C) Sanger sequencing results following SSO transfection, using primers specific for the transcript containing the pseudoexon, to confirm presence or absence of the wild-type c.2195C allele. (D) qPCR after SSO transfection, using primers specific for the transcript containing the pseudoexon. (E) qPCR after SSO transfection, using primers distant from the variants and amplifying total *GFM2* levels. Abbreviations: SSO – splice-switching oligonucleotides

Treatment of affected fibroblasts with SSO1 and SSO2 at two concentrations, 30pM and 150pM, revealed that SSO1 at 150pM was the most effective (Fig. S4). SSO1 (150pM) nearly abolished pseudoexon inclusion and restored the wild-type *GFM2* transcript (Fig. 2B and S4); subsequent experiments were therefore conducted using SSO1 at this concentration. Splicing correction was further confirmed by multiple complementary approaches in RNA extracted from treated fibroblasts. RT-PCR with primers amplifying both transcript isoforms, followed by gel electrophoresis, demonstrated a reduction in the transcript containing the pseudoexon. qPCR using primers specific for the pseudoexon-containing transcript and primers amplifying total *GFM2* transcript demonstrated a corresponding decrease in the aberrant transcript and an increase in total *GFM2* transcript levels (Fig. 2D,E).

Unexpectedly, the sequence-scrambled SSO increased the aberrant transcript (Fig. 2B). Although scrambled SSOs are commonly used as negative controls, antisense oligonucleotides can produce hybridization-dependent or chemistry-related off-target effects on RNA processing and transcript abundance (28). Similar nonspecific effects on poison-exon inclusion and NMD-sensitive transcripts have been reported (29). This observation underscores the importance of including mock-treated and chemistry-matched controls when interpreting SSO rescue experiments.

Having established that the pseudoexon-containing transcript was reduced after SSO treatment, we next sought to investigate whether this reduction reflected degradation of the pseudoexon transcript or genuine rescue through production of correctly spliced mRNA derived from it. To address this, we sequenced the transcript lacking the pseudoexon using an allele-specific forward primer that annealed to the wild-type junction between exons 19 and 20 (F_GFM2_Jx19-20) and a reverse primer on exon 21 (R_GFM2_Ex21) (Fig. 2A, S1), and evaluated the nucleotide present at the site of the point mutation in exon 20 (c.2195C>T), which is carried only by the non-duplicated allele. In untreated cells, all transcripts lacking the pseudoexon harbored the point mutation, and Sanger sequencing therefore showed only a thymine peak at this position. In SSO-treated cells, a cytosine peak reappeared at approximately comparable levels, indicating that a correctly spliced transcripts were also produced from the duplication-containing allele (Fig. 2C).

### SSO leads to increased protein levels and improved mitochondrial function

Following the observed transcript rescue, we assessed whether SSO1 increased mtEFG2 protein levels in affected and control fibroblasts, including cells transfected with the scrambled SSO. Although a ∼50% reduction in mtEFG2 protein levels may have been expected, given that one allele carries a missense variant predicted to produce a stable though dysfunctional protein, mtEFG2 was nearly undetectable in affected fibroblasts. This finding suggests that the missense variant may also destabilize the protein. mtEFG2 levels were partially restored following SSO1 transfection (Fig. 3A-C and S5).

**Figure 3.**
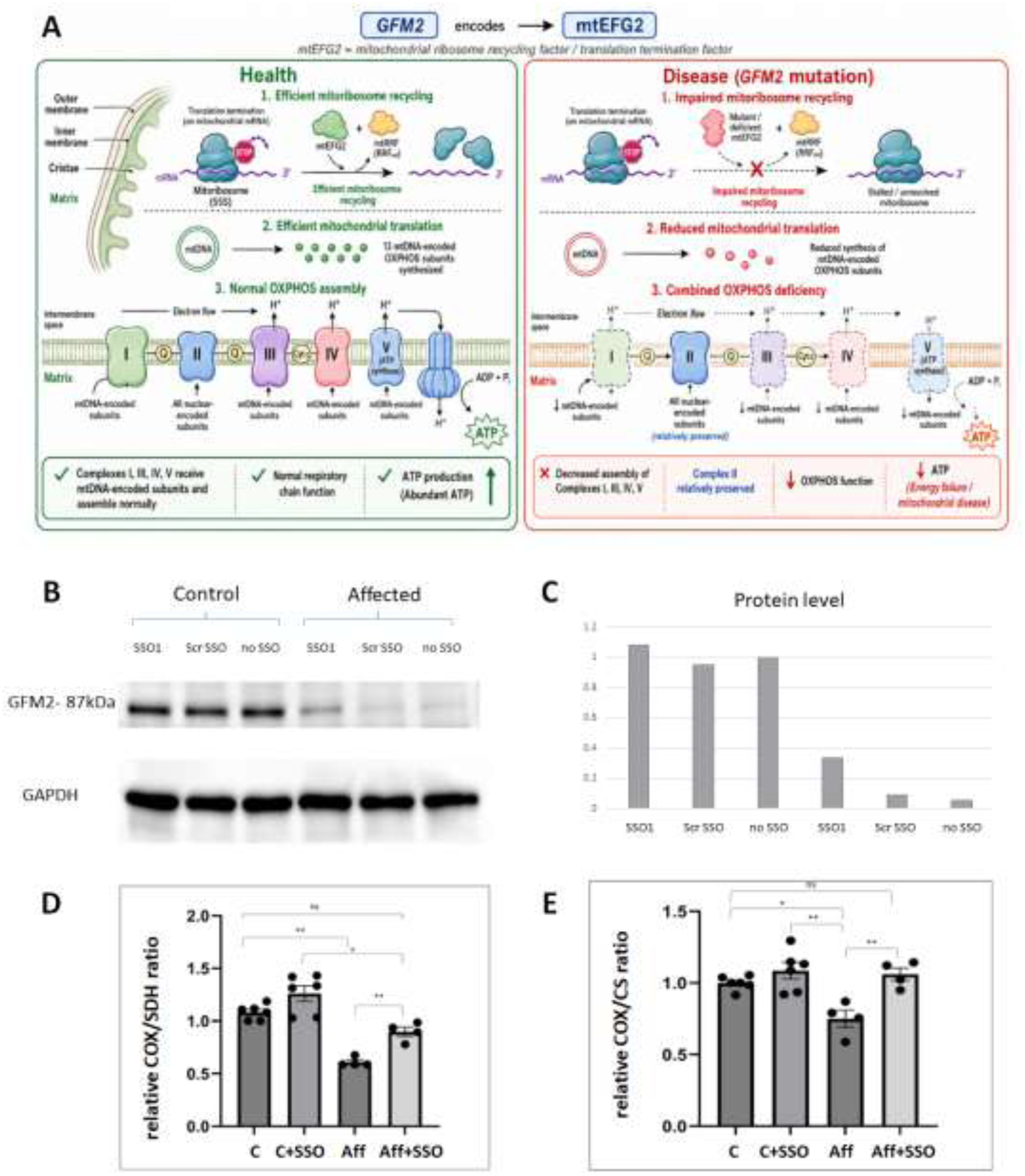
Protein levels and COX enzymatic activity ratios in affected fibroblasts with and without SSO treatment. (A) Schematic diagram showing the contribution of mtEFG2 (GFM2) activity to subunits of the electron transcript chain in health and disease (image generated according to authors’ prompt by Open AI’s ChatGPT, April 2026). (B) Western blot showing mtEFG2 protein levels in fibroblast cells from affected and a healthy control, with and without SSO transfection (150 pM). (C) Quantification of western blot protein levels using imageJ. (D) COX/SDH activity ratio. (E) COX/CS activity ratio. Abbreviations: COX – cytochrome C oxidase; CS – citrate synthase; SDH – succinate dehydrogenase

We then performed functional assays to determine whether increased mtEFG2 was accompanied by improved mitochondrial function. Since mtEFG2 is required for mitochondrial translation, we focused on COX activity. COX is one of three mtDNA-encoded subunits in the catalytic core of complex IV, and is expected to be sensitive to defects in mitochondrial translation. COX activity was reduced in affected fibroblasts (75.2 mU/mg) as compared with controls (107.9 ± 14.5 mU/mg [SEM]). The reduction was significant when COX activity was normalized to the nuclear-encoded complex II enzyme SDH and to the Krebs-cycle control enzyme citrate synthase (CS), corresponding to reductions of 39% and 25%, respectively (Fig. 3D,E). After 72 hours of SSO1 treatment, COX activity improved significantly to 105.7 mU/mg, reaching the control range. COX activity normalized to SDH and CS also improved significantly (to 147% and 141% of untreated fibroblasts, respectively), indicating functional respiratory-chain rescue (Fig. 3A-C). In contrast, SDH (38.9 mU/mg; controls 35.8 ± 4.5 mU/mg [SEM]) and CS (49.7 mU/mg; controls 52.8 ± 5.5 mU/mg [SEM]) activities were in the normal range and were not significantly altered by SSO1 treatment.

## DISCUSSION

We report an infant with a severe mitochondrial disorder caused by compound heterozygous *GFM2* variants: a coding missense variant and a 221-bp intronic duplication. RNA analysis showed that the duplication created the local sequence context needed for pseudoexon inclusion, generating an out-of-frame transcript targeted by NMD. The resulting reduction in mtEFG2 protein and selective impairment of the mtDNA-dependent COX activity support a loss-of-function mechanism consistent with defective mitochondrial translation. The study expands the molecular spectrum of *GFM2*-related disease by identifying an intronic structural variant that is pathogenic through splice activation rather than through disruption of coding sequence, and supports the value of integrated genome and transcriptome analyses in individuals with suspected mitochondrial disease, particularly when clinical and biochemical evidence strongly suggests a monogenic disorder but exome sequencing does not identify a complete explanation (6).

The diagnostic journey in this case is illustrative of the evolving role of multi-omic sequencing in mitochondrial disease. Exome sequencing has been shown to yield a definitive or possible molecular diagnosis in up to 60% of individuals with suspected mitochondrial disease in well-phenotype cohorts (30–32). However, while exome sequencing remains the first-tier test in many centers, its inherent limitations include poor coverage of deep intronic regions and structural variants. Although coding variants are enriched among known pathogenic alleles, intronic variants are an important and likely underrecognized cause of Mendelian diseases. Deep intronic variants can create or strengthen splice acceptor or donor sites, alter branch points or polypyrimidine tracts, disrupt regulatory motifs, or activate cryptic exons (33–36). Pseudoexon inclusion is especially damaging when it introduces a PTC, as in this case, because the mutant transcript is often degraded by NMD and the disease mechanism becomes functional haploinsufficiency or loss of function. The CHX experiment and allele-specific sequencing after SSO treatment were therefore central to showing both the fate of the aberrant transcript and the origin of the rescued wild-type transcript.

Most reported pseudoexon-activating variants are SNVs, but intronic insertions and duplications can have similar effects, as illustrated by several reported intronic duplications and insertions activating pseudoexons in *DMD* (37, 38). Because the importance of each nucleotide varies within the splice site sequence, changes at the most critical positions have the strongest impact and are more likely to overcome the repressive elements that normally prevent splicing at incorrect sites (35). This observation highlights why structural variants that are intronic by genomic position should not be presumed benign, particularly when they occur in a gene with a compelling phenotype match.

The development of molecular personalized therapies faces multiple challenges, including limited patient populations, limited data, high costs, and complex regulatory requirements. Traditional randomized control trials are often challenging, leading to consideration of alternative approaches such as N-of-1 therapies, where treatments are personalized for a single affected individual lacking other options. Antisense oligonucleotide (ASO) therapy has emerged as a promising strategy in this context, with a growing number of ASOs gaining approval and clinical trial activity, offering hope for personalized treatments in rare diseases (39–42).

Studies such as the present one can contribute to the emerging framework of individualized, mutation-specific therapies for rare genetic disorders (41,42). To our knowledge, this report represents one of the first demonstrations of SSO-mediated splicing correction leading to functional mitochondrial respiratory chain rescue in a primary mitochondrial disease caused by a nuclear-encoded mitochondrial translation factor. An analogous proof-of-concept study has been reported for another nuclear gene affecting mitochondrial function: Kumar et al. (2022) demonstrated SSO-mediated restoration of complex I subunit abundance and mitochondrial function in patient fibroblasts harboring a deep intronic *TIMMDC1* variant (43). Together, these reports position SSO therapy as a broadly applicable strategy for nuclear-encoded mitochondrial disease caused by splicing-disrupting variants.

SSO-based approaches are particularly tractable because: (i) the same chemical modification strategy (2’OMe/PS backbone) can be rapidly applied to new individual-specific splice variants;

(ii) SSOs do not require viral delivery or gene editing, reducing manufacturing complexity; and
(iii) the regulatory pathway for individualized ASO therapies (N-of-1) is increasingly defined, with frameworks now proposed by the Food and Drug Administration (FDA) for compassionate treatments.(44) Seminal clinical examples include milasen for *CLN7*-related Batten disease (45) and individualized ASOs for *SCN8A* epilepsy (46), demonstrating proof-of-principle in pediatric neurological diseases analogous to the present case.

A critical prerequisite for this translational step is the type of *in-vitro* validation presented here: demonstrating transcript correction, protein rescue, and functional improvement in patient-derived cells, before committing to clinical-grade synthesis and delivery optimization. However, this study has several important limitations. First, fibroblasts were used as the cellular model; although they are accessible and informative for mitochondrial disorders, they may not fully recapitulate the tissue specificity of *GFM2*-related pathology, particularly in neuronal or muscle tissues where mitochondrial translation deficits may be more pronounced. Second, the lack of an *in vivo* model precludes assessment of tissue distribution, pharmacokinetics, toxicity, and long-term efficacy of the SSO. These factors are critical for evaluating therapeutic feasibility beyond the cellular level.

Completion of the preclinical trials would require lead optimization, off-target assessment, biodistribution and pharmacokinetic studies, and toxicology in rodent models. In addition, studies in iPSC-derived neurons or organoid models could help determine whether rescue is reproducible in more relevant cellular systems, and may define a therapeutically meaningful threshold of mtEFG2 restoration. The observation that the scrambled SSO altered the abundance of the aberrant transcript further supports the need for orthogonal controls and systematic off-target evaluation, particularly because hybridization-mediated off-target effects have been described for splice-switching ASOs (28).

## CONCLUSIONS

We identify a pathogenic intronic duplication in *GFM2* that activates a cryptic splice site and leads to deleterious pseudoexon inclusion. We further demonstrate that an SSO designed to block this aberrant splice junction restores correct mRNA processing, increases mtEFG2 protein, and improves mitochondrial respiratory-chain function in affected fibroblasts. These findings demonstrate how integrated DNA–RNA diagnostics can reveal actionable mechanisms in unresolved mitochondrial disease and support splice-switching therapy as a promising, mechanism-based approach for selected rare genetic disorders caused by aberrant splicing.

## LIST OF ABBREVIATIONS

2’OMe: 2’-O-methyl
ASO: antisense oligonucleotide
ATP: adenosine triphosphate
CNV: copy number variants
COX: cytochrome c oxidase
CS: citrate synthase
CSF: cerebrospinal fluid
ETC: electron transport chain
IDT: integrated DNA technologies
MAF: minor allele frequency
MRI: magnetic resonance imaging
mtDNA: mitochondrial
DNA NMD: nonsense mediated decay
OXPHOS: oxidative phosphorylation
PTC: premature termination codon
qPCR: quantitative polymerase chain reaction
SNV: single nucleotide variants
SSO: splice-switching oligonucleotide
SDH: succinate dehydrogenase
WGS: Whole genome sequencing

## DECLARATIONS

### Ethics approval and consent to participate

The family was consented for research studies by written informed consent, according to Helsinki committee-approved protocol 0306-10-HMO.

### Consent for publication

The parents provided written consent for publication.

### Availability of data and materials

The variant data has been submitted to ClinVar (Variation ID: 4755559 and 4755560).

### Competing Interests

H.M.S is an employee of Geneyx Genomix. Other authors declare no competing interests.

## Funding

This study was supported in part by The Estate of David Weinstock (TH).

## Authors’ Contributions

SG and TH conceived and designed the molecular studies; RB and HMS interpreted genomic and transcriptomic data; NSL and JM contributed detailed clinical data; SG, CR and ASh performed RNA studies; SG and SYD conducted molecular assays; ASa, SG and ALM conducted enzymatic and statistical analyses; SG and TH wrote the draft and all authors revised, edited and approved the final submitted version.

## Data Availability

The variant data has been submitted to ClinVar (Variation ID: 4755559 and 4755560).

https://www.ncbi.nlm.nih.gov/clinvar/variation/4755560/?term=4755560%5BVariation+ID%5D

## Acknowledgements

The authors thank the family for participation in this study.

## Notes

### Author Declarations

The research study was approved by the Hadassah Medical Center institutional review board. The family was consented for research studies by written informed consent, according to Helsinki committee-approved protocol 0306-10-HMO.

