## Supplemental Tables and Figures for "In vitro splice-switching oligonucleotide rescues aberrant *GFM2* pseudoexon inclusion and restores mitochondrial activity"

**Table S1. Primers used in the study**

| Primer name | Seq (5'->3') |
|---|---|
| F_GFM2_int19 | GATTTCAAAGGCCAAGAAGAACA |
| R_GFM2_int19 | GATGAGCTCAGTGTGGGACA |
| F_GFM2_Ex16 | TCAGCGTGAAGATCCCAGTT |
| R_GFM2_Ex17 | TGGTCTCTCGATATGCCACC |
| F_GFM2_Jx18-19 | GTCTCCAAGGACCATTGCT |
| F_GFM2_Ex19 | CAATTCATCCTGGCACCTCC |
| F_GFM2_Jx19-20 | GCGTGCAAAAGGCTCTGAAG |
| F_GFM2_Jx19-PE | GCAAAAGGACTGGAGCACTC |
| R_GFM2_Ex20 | CTTTGTTGTCCTGGCGAGTC |
| R_GFM2_Ex21 | GTAGCTGAGCCTGATGTTAGC |

**Table S2. Splice-switching oligonucleotides**

| SSO | Seq (5'->3') |
|---|---|
| SSO1 | GUGCUCCAGUCCUGUUGAAAAGG |
| SSO2 | CAAUAUUACCUAUCUGGAGAGC |
| ScrSSO | CCAGUGUGCUAAGGUGAACCUGU |

**Abbreviations:** SSO – splice-switching oligonucleotide; ScrSSO – scrambled SSO

### Table S3. Clinical features of individuals with biallelic pathogenic or likely pathogenic variants

| Feature | Individual 1 | Individual 2 | Individual 3a-b | Individual 4 | Individual 5 | Individual 6 | Individual 7 | Total |
|---|---|---|---|---|---|---|---|---|
| **Reported individual(s)** | Fukumura et al., 2015 (Sibling 1) | Fukumura et al., 2015 (Sibling 2) | Dixon-Salazar et al., 2012 (Affected siblings/ Family 650) | Glasgow et al., 2017 (Individual 1) | Glasgow et al., 2017 (Individual 2) | Gouiza et al., 2024 | Current study | |
| **Gender** | Female | Female | | Male | Female | Female | Male | |
| **Age at presentation** | Birth | Birth | Neonatal | 2y 6mo; regression from 5y | 2y 2mo | 1y | Birth | |
| **Neurological phenotype** | | | | | | | | 7/7 |
| **Severe global developmental delay (HP:0001263)** | + | + | + | + | + | + | + | |
| **Developmental regression (HP:0002376)** | | | | + | + | | | |
| **Epilepsy/seizures (HP:0001250)** | + | - | | | + | + | + | |
| **Hypotonia (HP:0001252)** | + | + | | | | | + | |
| **Microcephaly (HP:0000252)** | | | + | | | | | |
| **Dysarthria (HP:0001260)** | | | | + | | | | |
| **Ocular phenotype** | | | | | | | | 3/7 |
| **Optic atrophy (HP:0000648)** | + | + | | | | | | |
| **Reduced eye contact (HP:0000817)** | + | + | | normal vision | | | + | |
| **Nystagmus (HP:0000639)** | | | | | | + | | |
| **Musculoskeletal / tone / movement phenotype** | | | | | | | | 5/7 |
| **Arthrogryposis multiplex congenita (HP:0002804)** | + | + | | | | | | |
| **Dystonia (HP:0001332)** | | | | + | + | + | | |
| **Spasticity (HP:0001257)** | | | | + | + | | | |
| **Other** | | | | | | | | |
| **Bradycardia (HP:0001662)** | + | + | | | | | | |
| **Insulin-dependent diabetes mellitus (HP:0000819)** | | | + | | | | | |
| **Hypospadias (HP:0000047), Cryptorchidism (HP:0000028), Inguinal hernia (HP:0000023)** | | | | | | | + | |
| **Biochemical findings** | | | | | | | | 6/7 |
| **Elevated lactate (HP:0003128)** | Lactate peak on MRS | Lactate peak on MRS | NA | High (CSF) | High (plasma and CSF) | High (plasma and CSF) | High (plasma and CSF) | |
| **ETC/ OXPHOS** | Low complex III and IV activities | Low complex III and IV activities | NA | Muscle complex IV deficiency; reduced mtEFG2 and COX I/II in muscle | Reduced steady-state OXPHOS subunits across complexes I, III, and IV by Western blot | | | |
| **Brain MRI** | | | | | | | | 7/7 |
| **Ventriculomegaly/enlarged lateral ventricles (HP:0002119)** | + | | | | | | | |
| **Hypoplasia of the corpus callosum (HP:0002079)** | + | | | | | | | |
| **Cerebral atrophy (HP:0002059)** | | + | | | | | | |
| **Simplified gyral pattern (HP:0009879)** | | | + | | | | | |
| **Cerebellar atrophy (HP:0001272)** | | + | | + | | | | |
| **Abnormal basal ganglia MRI signal (HP:0012751)** | | | | + | + | + | + | |
| **Corpus callosum abnormality (HP:0001273)** | | | | + | + | + | | |
| **Subcortical white-matter involvement (HP:0002500)** | | | | | + | + | | |
| ***GFM2* variant(s)** | | | | | | | | |
| **Variant/s (NM_032380.5)** | c.206+4A>G / c.2029-1G>A - compound heterozygous | c.206+4A>G / c.2029-1G>A - compound heterozygous | c.1728T>A, p.(Asp576Glu) - homozygous (splice-altering) | c.569G>A, p.(Arg190Gln) / c.636delA, p.(Glu213Argfs*3) - compound heterozygous | c.275A>C, p.(Tyr92Ser) - homozygous. | c.569G>A, p.(Arg190Gln) - homozygous | c.2195C>T, p.(Pro732Leu) / c.2029-741_2029-521dup - compound heterozygous | |

**Abbreviations:** CSF, cerebrospinal fluid; ETC, electron transport chain; HP, Human Phenotype Ontology; MRI, magnetic resonance imaging; MRS, magnetic resonance spectroscopy; NA, not available; OXPHOS, oxidative phosphorylation

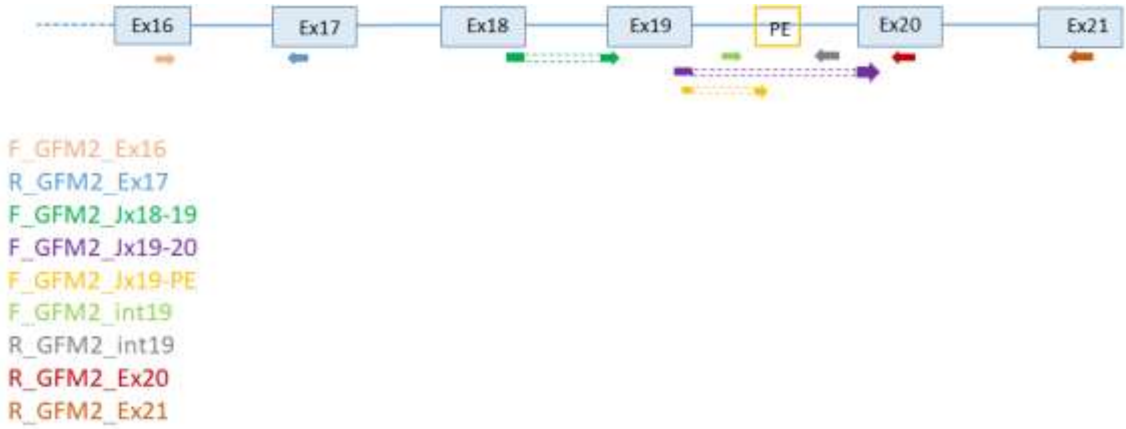


**Figure S1. Schematic diagram of primers used in the study.**

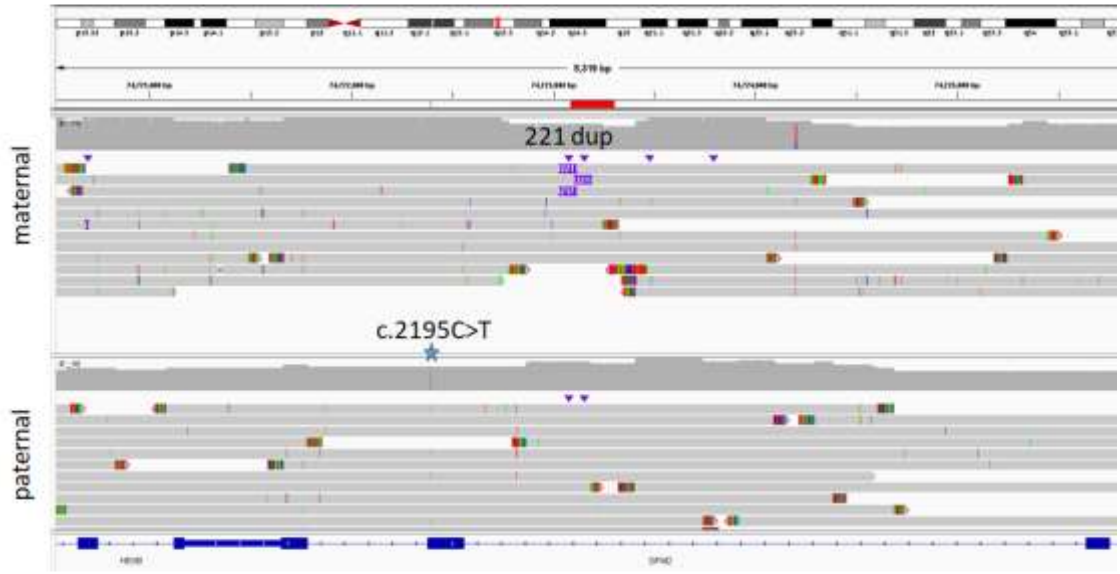


**Figure S2. Long read sequencing of parental DNA.** Upper panel shows heterozygous 221 duplication in maternal DNA. Lower panel shows heterozygous c.2195C>T variant. Note that gene is transcribed from the minus strand.

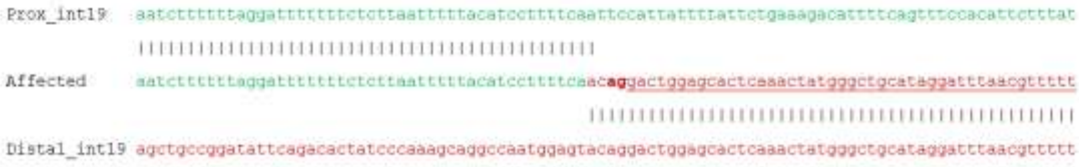


**Figure S3. Breakpoint junction sequence.** Middle sequence indicates affected individual, whereas top (green) and bottom (red) sequences show proximal and distal sequences in intron 19. Underlined sequence represents pseudo-exon inclusion at the RNA level, and letters in bold (**ag**) indicate activated cryptic acceptor splice site. Polypyrimidine tract rich in thymine (T) and cytosine (C) nucleotides, positioned before the cryptic splice site in the affected individual, can be appreciated.

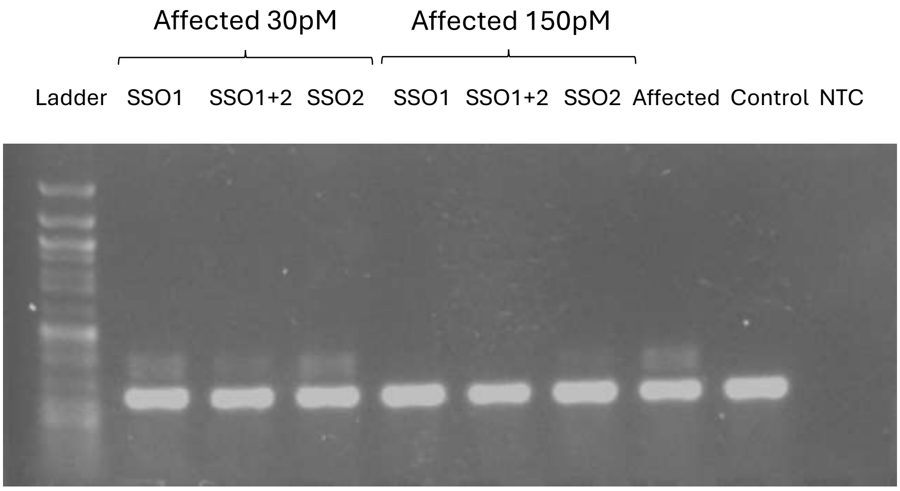


**Figure S4. Effect of SSO1 and SSO2 at two concentrations.** The effect of SSO1 at 150pM abolished the pseudoexon.

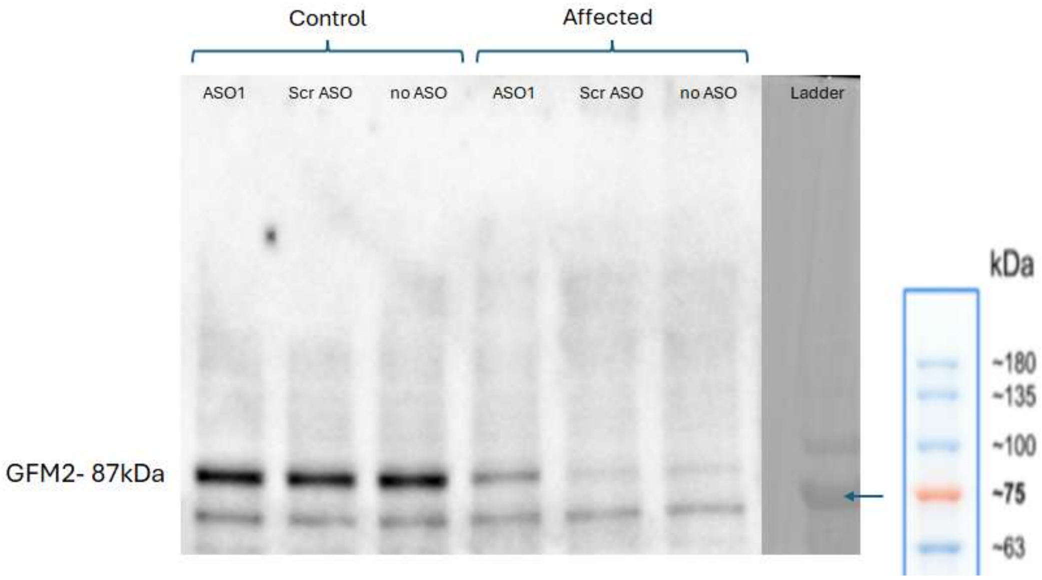


**Figure S5. Western blot.** Western blot (uncropped) indicating reduced GFM2 protein levels in affected cells, with partial rescue by ASO1.
